# Two artificial tears outbreak-associated cases of XDR *Pseudomonas aeruginosa* detected through whole genome sequencing-based surveillance

**DOI:** 10.1101/2023.04.11.23288417

**Authors:** Alexander J. Sundermann, Vatsala Rangachar Srinivasa, Emma G. Mills, Marissa P. Griffith, Kady D. Waggle, Ashley M. Ayres, Lora Pless, Graham M. Snyder, Lee H. Harrison, Daria Van Tyne

## Abstract

We describe two cases of XDR *Pseudomonas aeruginosa* infection caused by a strain of public health concern recently associated with a nationwide outbreak of contaminated artificial tears. Both cases were detected through database review of genomes in the Enhanced Detection System for Hospital-Associated Transmission (EDS-HAT), a routine genome sequencing-based surveillance program. We generated a high-quality reference genome for the outbreak strain from one of the case isolates from our center and examined the mobile elements encoding *bla*_VIM-80_ and *bla*_GES-9_ carbapenemases. We then used publicly available *P. aeruginosa* genomes to explore the genetic relatedness and antimicrobial resistance genes of the outbreak strain.

## INTRODUCTION

On February 1, 2023, the Centers for Disease Control and Prevention (CDC) released a Health Alert Network health advisory for an outbreak of Verona Integron-encoded Metallo-β-lactamase (VIM) and Guiana Extended-Spectrum β-Lactamase (GES)-producing carbapenem-resistant *Pseudomonas aeruginosa* (VIM-GES-CRPA) associated with the use of artificial tears.^1^ As of March 2023, there have been 68 cases detected among specimens collected from May 2022 to February 2023 in 16 states, including three deaths and reports of local transmission within healthcare facilities.^2,3^ The specific VIM-GES-CRPA strain belongs to multi-locus sequence type (ST) 1203 and contains resistance genes *bla*_VIM-80_ and *bla*_GES-9_.^2^ Before this outbreak, such a strain of VIM-GES-CRPA had not been detected in the United States.

The CDC has encouraged healthcare facilities to report possible cases of carbapenemase-producing *P. aeruginosa* with epidemiological risk factors of exposure to presumably contaminated eye drop products. More recently, the CDC provided genome sequences of representative outbreak isolates for healthcare facilities and laboratory scientists to compare available genomes against the outbreak strain.^2^ Thus, detection relies on astute clinical investigation and/or a healthcare facilities’ whole genome sequencing (WGS) and analysis capabilities.

Here, we describe our experience identifying the VIM-GES-CRPA outbreak strain at our facility by leveraging a WGS surveillance and control program called the Enhanced Detection System for Healthcare-Associated Transmission (EDS-HAT).^4–9^ Using available representative outbreak isolate genomes, we identified three previously collected and sequenced *P. aeruginosa* isolates sampled from two patients at our center that were highly related to each other and to the outbreak isolates. We generated a hybrid assembled reference sequence for an outbreak isolate from our hospital, explored the carbapenemase-encoding mobile elements present in the strain, and constructed a global phylogeny of ST1203 clinical isolates, which included 50 putative outbreak isolates collected from the United States in the last 18 months.

## METHODS

### Study setting

The University of Pittsburgh Medical Center-Presbyterian Hospital (UPMC) is an adult tertiary care hospital with 758 total beds (including 134 critical care beds) and performs over 400 solid organ transplants annually. The Institutional Review Board of the University of Pittsburgh gave ethical approval for this work under Protocol STUDY21040126.

### Isolate collection & WGS

Real-time WGS surveillance from EDS-HAT was initiated in November 2021 for hospital-associated pathogens of high concern, as previously described.^4^ Briefly, twice per week potentially healthcare-associated clinical cultures, as defined by a hospital stay ≥3 days or a recent healthcare exposure in the prior 30-days, were collected. *P. aeruginosa* was included in this collection. Genomic DNA was extracted from all isolates and sequenced weekly on the Illumina platform.^4^ When warranted, long read sequencing of select isolates was performed on the Oxford Nanopore MinION platform.^10^ Hybrid assembly of Illumina short reads and Nanopore long reads was performed with unicycler to resolve mobile genetic elements.^11^ *P. aeruginosa* isolates were considered part of a genetically related cluster if ≥2 isolates differed from one another by no more than 15 single nucleotide polymorphisms (SNPs) using split kmer analysis (ska).^4,12^ Additional *P. aeruginosa* ST1203 genomes, including those of three representative outbreak isolates, were downloaded from the NCBI database on April 6, 2023 (Table S1). A phylogenetic tree was made using RAxML from a core genome alignment generated with snippy (https://github.com/tseemann/snippy).^13^ Antimicrobial resistance (AMR) genes were identified with AMRFinderPlus.^14^ Integrative and conjugative elements (ICEs) were identified using ICEFinder https://bioinfo-mml.sjtu.edu.cn/ICEfinder/ICEfinder.html).

### Infection Prevention and Control (IP&C)

Clusters of genetically related bacteria were initially investigated by the research team and findings were shared weekly with the IP&C Department. The IP&C Department performed an additional investigation for possible epidemiological links within clusters and initiated appropriate interventions aimed at halting transmission. Clusters were monitored regardless of epidemiological links, and if additional clinical cultures were identified then additional investigations were performed.

## RESULTS

From November 2021 through March 2023, we collected and sequenced 867 *P. aeruginosa* isolates as part of the EDS-HAT program. Three (0.35%) isolates collected from two patients in October 2022 were identified as belonging to ST1203. These isolates formed a WGS cluster with each other (differing by 0-4 SNPs); however, initial investigation revealed no common exposures between the two patients. Patient 1 had two cultures from ear drainage as an outpatient presenting with complaints of an ear infection starting in early 2022, while Patient 2 had an isolate recovered from a bronchoalveolar lavage during a stay in an intensive care unit in October 2022.

In April 2023, after reviewing updates on the VIM-GES-CRPA artificial tears outbreak from the CDC including information about the ST of the outbreak strain and associated AMR genes, we retrospectively examined our WGS database for ST1203 *P. aeruginosa* isolates and evaluated these for the presence of *bla*_VIM-80_ and *bla*_GES-9_. We identified two isolates from Patient 1 and one isolate from Patient 2 that belonged to ST1203 and encoded both carbapenemase genes at 100% identity. We also identified an additional ST1203 isolate in our EDS-HAT retrospective data set^4^ collected in 2019 that did not encode either carbapenemase gene. Using the available representative isolate genomes from the CDC, we found that the three isolates from 2022 were closely related to one another as well as to the representative outbreak isolates (0-7 SNPs), while the retrospective 2019 isolate was unrelated (>780 SNPs) (Figure 1A). The IP&C Department and public health partners were immediately notified. Additional chart review for Patient 1 revealed that the patient had been using eye drops purchased from an online retailer before the presentation of the initial complaint in early 2022. Review of Patient 2 chart records did not reveal any apparent over-the-counter eye drop use. No epidemiological commonalities between the two patients were found.

**Figure 1.**
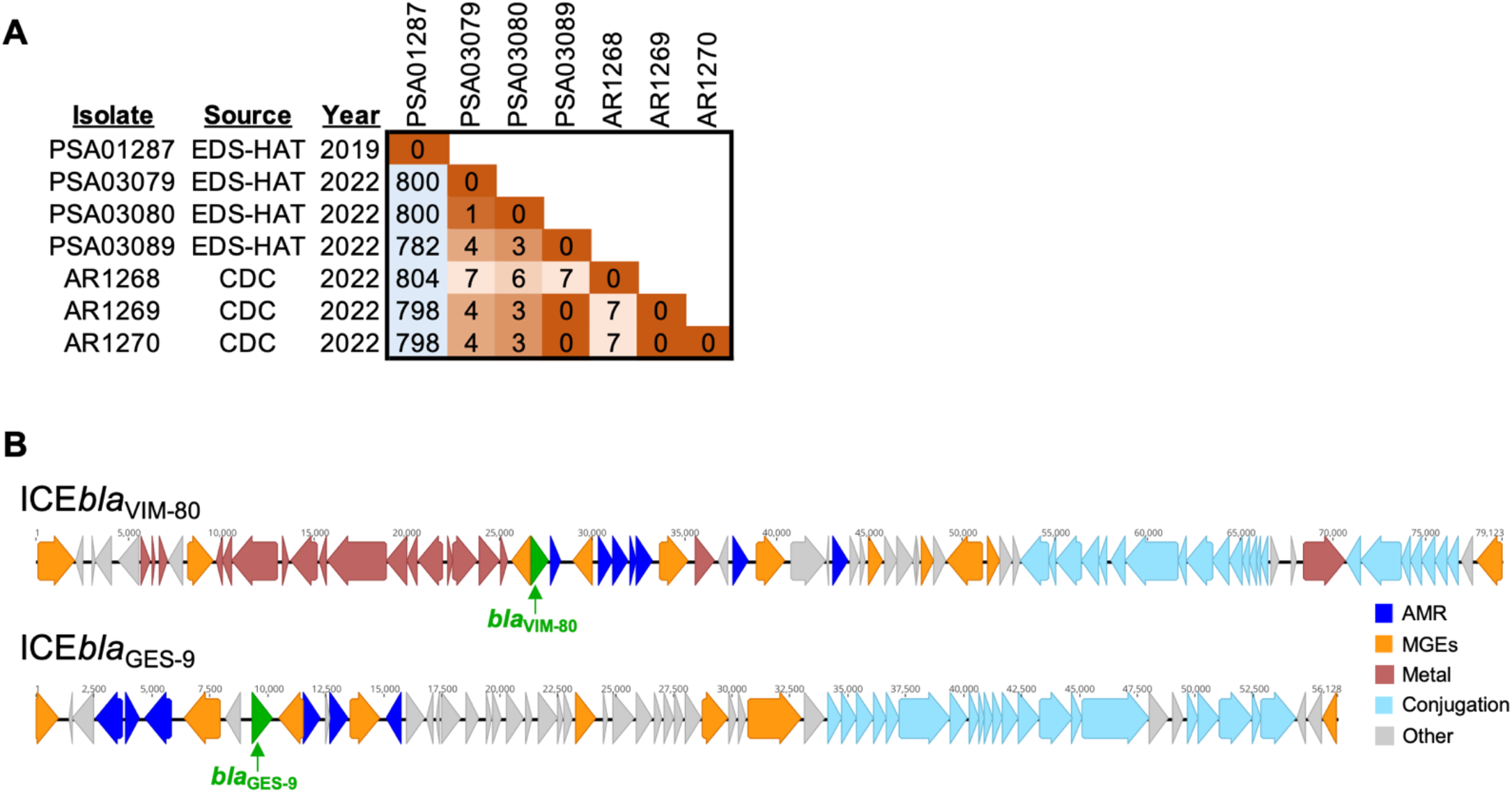
Identification of outbreak-associated isolates and carbapenemase-encoding mobile genetic elements. (A) Pairwise single nucleotide polymorphism (SNP) matrix for four ST1203 isolates collected through EDS-HAT (PSA01287, PSA03079, PSA03080, PSA03089) and three representative outbreak isolates provided by CDC (AR1268, AR1269, AR1270). SNPs were identified with split kmer analysis (ska), and matrix colors correspond to SNP distance between isolates. (B) Integrative conjugative elements (ICEs) carrying *bla*_VIM-80_ and *bla*_*GES*-9_ in isolate PSA03079. ICEs were identified with ICEFinder and were extracted from the hybrid genome assembly of PSA03079. Coding sequences are colored by putative function. Green = carbapenemases; dark blue = antimicrobial resistance (AMR) genes; orange = mobile genetic element genes (MGEs); red = metal-interacting genes (Metal); light blue = conjugation machinery genes (Conjugation); grey = other.

The genomic regions encoding the *bla*_VIM-80_ and *bla*_GES-9_ carbapenemases were initially unresolved in draft genome assemblies constructed from Illumina short-read data. We performed long-read sequencing and hybrid assembly of PSA03079 (collected from Patient 1) and generated a high-quality draft genome assembly composed of eight contigs. We identified four different regions in the hybrid assembled genome as putative integrative and conjugative elements (ICEs). One of these ICEs encoded the *bla*_VIM-80_ carbapenemase as well as genes involved in heavy metal resistance (Figure 1B). A second ICE encoded the *bla*_GES-9_ carbapenemase and the aminoglycoside resistance gene *rmtF*, and resembled the previously described ICE*6660*.^15^ The sequences of both ICEs resembled similar elements found in other publicly available genomes; however, both carbapenemases appeared to have been inserted into these elements via additional, smaller mobile genetic elements. These data suggest that both carbapenemases are mobilizable and could be transmitted to other pathogens through horizontal gene transfer. We searched all genomes in the EDS-HAT database for *bla*_VIM-80_ and *bla*_GES-9_ and did not find any additional isolates that encoded them at the time of manuscript submission.

We next queried the NCBI database for additional genomes of *P. aeruginosa* isolates belonging to ST1203. We selected isolates from clinical sources and with known dates of isolation. Genomes were downloaded and were combined with EDS-HAT and CDC representative isolates to construct a global phylogeny of 69 isolates belonging to the ST1203 lineage (Figure 2). The phylogeny showed that 47 isolates closely related to the representative outbreak isolates provided by CDC were collected from the United States in 2022 and 2023, including the three isolates sampled by EDS-HAT. Additionally, we found that the outbreak strain was most closely related to ST1203 isolates collected between 2013 and 2018 from India and Nigeria (Figure 2). These earlier isolates encoded *bla*_GES-9_ but not *bla*_VIM-80_, suggesting independent acquisition of each carbapenemase. A variety of other AMR genes were detected among ST1203 isolates; however, isolates belonging to the outbreak strain had similar AMR gene profiles. Taken together, these data suggest that ST1203 was a rarely observed lineage prior to 2022, and that many outbreak strains have been sampled and sequenced in the United States since the reported start of the VIM-GES-CRPA artificial tears outbreak.

**Figure 2.**
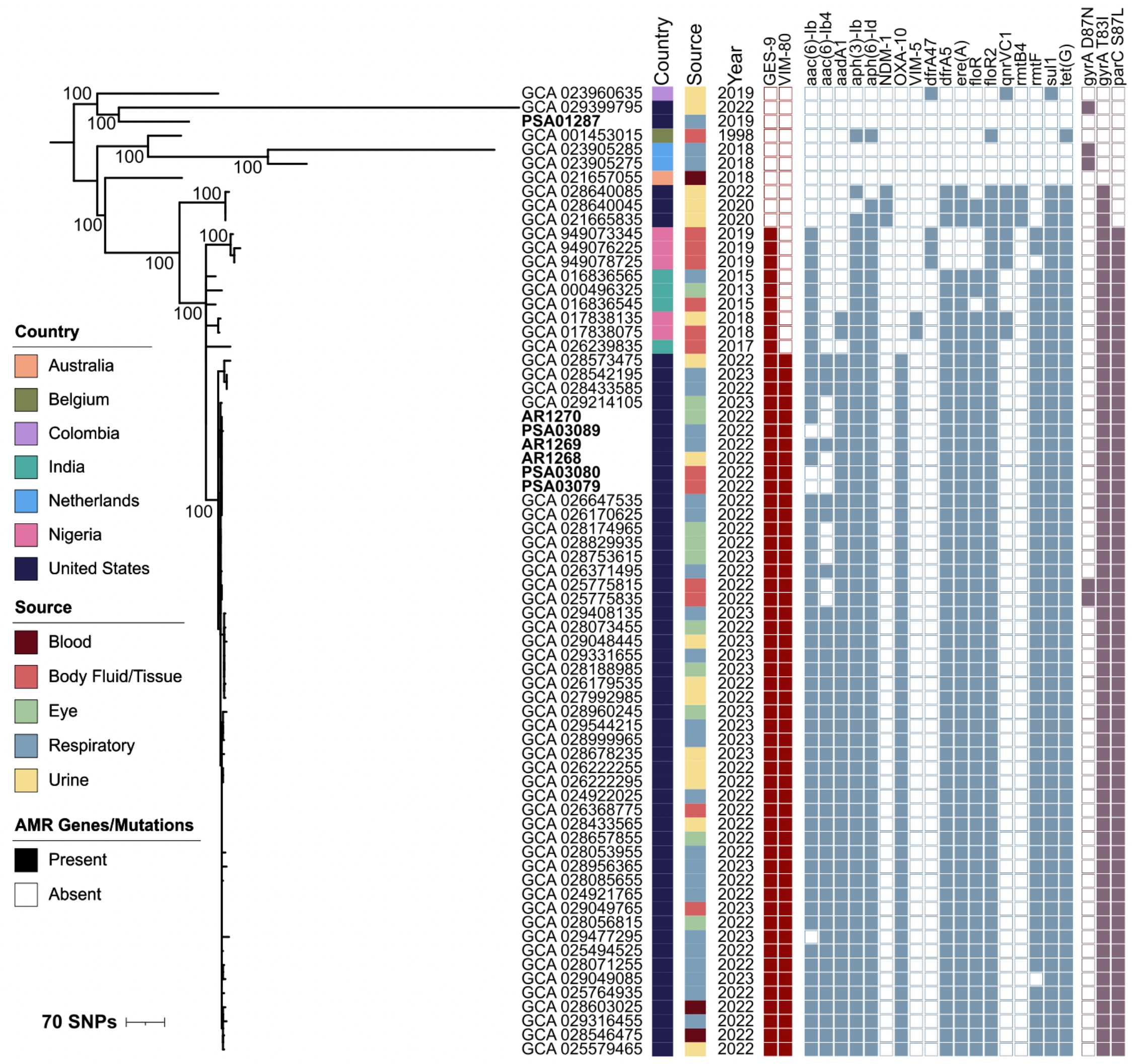
Global phylogeny of publicly available ST1203 *P. aeruginosa* genomes. Genome assemblies were downloaded from NCBI and a core genome phylogeny was constructed with snippy and RAxML. The phylogeny was annotated with the country, source, year of isolation, and AMR genes and mutations identified in each isolate. Bootstrap values are listed next to nodes with high confidence support.

## DISCUSSION

In this report, we describe the detection of two cases of VIM-GES-CRPA putatively belonging to the national outbreak associated with the use of artificial tears through EDS-HAT, an ongoing sequencing-based surveillance program at our institution. Detection of this outbreak strain was only possible through the open data sharing of representative outbreak isolate genomes and strain-identifying genetic information provided by the CDC. EDS-HAT enabled the IP&C Department to rapidly investigate, evaluate, and ensure that no additional transmission occurred, and to promptly notify public health officials of additional cases related to the ongoing outbreak.

Our findings and investigation show the value of healthcare pathogen WGS surveillance programs for outbreak detection and intervention. We previously described the infection prevention implications of initiating such a program, including uncovering new outbreaks, interrupting transmission, enhancing patient safety, and generating significant cost savings.^4,5,7–9,16^ EDS-HAT detects outbreaks and transmission at our facility as soon as two patients, allowing for prompt intervention.^6^ If instituted broadly at other healthcare facilities with open data sharing, outbreaks such as the VIM-GES-CRPA from artificial tears could be more rapidly detected, given the nature of medication contaminated outbreaks. Detection triggers of multi-state or facility outbreaks often require a unique pathogen, as the one described here was not previously detected in the United States, or an incidence high enough for public health notification and investigation.

There are several limitations to this study. First, we only performed WGS on isolates from patients with clinical infections and specific pathogens who had recent healthcare exposure. Given the nature of this outbreak, we could have missed additional cases if patients did not have recent healthcare exposure. Second, we only detected these isolates retrospectively once WGS data from representative outbreak isolates were available. Finally, although we did not detect any evidence of onward transmission at our institution, we did not look for possible transmission to asymptomatic colonized individuals. Ongoing screening initiatives for exposed individuals is underway.

In conclusion, we describe the detection of two cases of VIM-GES-CRPA associated with the national outbreak from artificial tears that we detected through an ongoing healthcare WGS surveillance program. As WGS becomes more widely available, we expect that rapid outbreak identification and subsequent case detection will become more common. Healthcare institutions should consider the adoption of WGS surveillance-based tools to prevent transmission of AMR pathogens and enhance patient safety.

## Data Availability

All data produced in the present study are available upon reasonable request to the authors.

## ACKNOWLEDGMENTS

We thank the UPMC IP&C Department and Vaughn Cooper for whole genome sequencing support.

